# Evaluation of a Rapid Implementation of Telemedicine for Delivery of Obstetric Care During the COVID-19 Pandemic

**DOI:** 10.1101/2021.05.19.21257311

**Authors:** Chinyere N. Reid, Jennifer Marshall, Kimberly Fryer

## Abstract

**Objective:** The aim of this evaluation was to assess the rapid implementation of obstetric telemedicine during the COVID-19 pandemic using the Consolidated Framework in Implementation Research (CFIR) evaluation framework.

**Study Design:** Following one month of telemedicine implementation, obstetric providers at the University of South Florida clinic completed qualitative surveys and in-depth interviews about the implementation of obstetric telemedicine in the clinic guided by the CFIR evaluation framework.

**Results:** Overall, providers considered obstetric telemedicine comparable to traditional in-person clinic visits and acknowledged that they were adequately prepared for the telemedicine implementation. Advantages included the simplicity of implementation, reduced exposure to COVID-19 infection, and convenience factors. Although obstetric telemedicine mostly met patient needs, a lack of access to at-home monitoring devices, physical examinations, reliable internet service, and privacy concerns posed as barriers.

**Conclusion:** The implementation of the obstetric telemedicine care model was deemed a favorable, safe alternative option for patients during the COVID-19 pandemic.

## Introduction

The 2019 novel coronavirus (COVID-19), originating in Wuhan, China, subsequently became a global pandemic infecting over 108 million people and resulting in over two million deaths to date.^1^ In the United States (US) more than 27 million people have tested positive for COVID-19 and over 470,000 deaths have occurred.^2^ In the initial stages of the COVID-19 pandemic, lockdowns, quarantines, and social distancing measures were implemented because of the high transmissibility of the COVID-19 virus, shortage of personal protective equipment, and the absence of curative treatments or a vaccine.

As a result, these measures impeded the provision and participation of traditional in-person healthcare appointments. For obstetric patients, the need to limit infection exposure, reduced access to childcare because of school closures, and restrictions to working from home were barriers likely to prevent access to in-person obstetric care. Likewise, in response to the COVID-19 pandemic, preventive social measures were likely to impact health-related policies and practices thereby limiting the delivery of comprehensive obstetric care services.^3^ In order to support the continuity of healthcare services during the COVID-19 pandemic while adhering to pandemic restrictions, there was an increase in the adoption and utilization of telemedicine for the delivery of healthcare services.

Telemedicine involves two-way audio and visual electronic communication between patients and providers in real-time at different remote locations.^4^ The American Hospital Association reported that in 2017, 76% of US hospitals connected patients remotely with healthcare providers using video or other technology.^5^ Research has also shown that prior to the COVID-19 pandemic, Radiology specialists had the highest utilization rates of telemedicine (39.5%), while Obstetrician-Gynecologist specialists had among the lowest utilization rates of telemedicine (9.3%) in their practices.^6^ Telemedicine has been shown to be advantageous for both patients and healthcare providers, due to its ease of accessibility to attend office visits from different geographical locations, reduced expenses related to travel costs and childcare costs, increased time efficiency for consultations, and limited exposure to infectious diseases.^7^

Although most studies have assessed the implementation of telemedicine in various clinical settings without the guidance of theory,^8–14^ we aimed to assess the rapid implementation, feasibility, and satisfaction of obstetric telemedicine during the COVID-19 pandemic using the Consolidated Framework in Implementation Research (CFIR) evaluation framework.^15^

## Subjects and Methods

### Overview

This qualitative evaluation was developed using CFIR, which provides constructs that have been associated with effective implementation.^16^ Key CFIR constructs guiding this evaluation included Intervention Characteristics (Evidence Strength and Quality, Relative Advantage, Adaptability, Complexity), Outer Setting (Patient Needs & Resources), Inner Setting (Implementation Climate, Readiness for Implementation), Characteristics of Individuals (Knowledge & Beliefs about the Intervention), and Processes (Engaging Intervention Participants) related to the implementation of obstetric telemedicine.

### Data Collection

We recruited obstetric providers delivering obstetric care visits via telemedicine at the University of South Florida clinic from May to November 2020. Purposive sampling was used to recruit obstetric providers via email who were provided with a link to complete an anonymous online survey. Eligibility criteria included healthcare providers that currently provided obstetric care at University of South Florida clinic. Nineteen participants completed the online survey out of 31 total prenatal care providers in the practice (Table 1).

**Table 1.** Consolidated Framework for Implementation Research (CFIR) constructs used to assess implementation of obstetric telemedicine visits guided by CFIR^*^

| Domain | Construct | Definition |
| --- | --- | --- |
| Intervention Characteristics | Evidence Strength and Quality | Perception about the quality and validity of evidence supporting the belief the intervention will have desired outcomes |
|  | Relative Advantage | Perception of the advantage of implementing the intervention compared to other programs |
|  | Adaptability | Degree to which intervention can be changed to meet local needs |
|  | Complexity | Perception about the difficulty of implementing the intervention |
| Outer Setting | Patient Needs and Resources | Extent to which the needs of patients are known and prioritized |
| Inner Setting | Implementation Climate | Shared receptivity of individuals to an intervention |
|  | Readiness for implementation | Actual organizational commitment to implement the intervention |
|  | Available Resources | Level of resources allocated for implementation and continued use of intervention |
|  | Access to Knowledge and<br>Information | Access and use of knowledge and<br>information about the intervention |
| Characteristics<br>of Individuals | Knowledge and Beliefs<br>about the Intervention | Individual's attitudes and values towards the<br>intervention |
| Process | Engaging | Encouragement of individuals in the<br>implementation and use of the intervention |

\*These CFIR items <sup>16</sup> were specifically intended for this study and constructs are listed in the order they appear in the text.

The anonymous online Qualtrics survey included a demographic questionnaire on providers’ age, race, and years in practice post residency or fellowship training, and open-ended questions guided by key CFIR constructs.^17^ We selected eight key CFIR constructs most relevant to implementation of obstetric telemedicine. Some open-ended survey questions included: *“What advantages do telemedicine visits have compared to existing programs/in-patient visits?”* (Intervention Characteristics); *“What barriers do you believe obstetric patients face in participating in telemedicine visits?”* (Outer Setting); *“What was the general level of receptivity in your clinic to implementing telemedicine visits?”* (Inner Setting); *“What steps were taken to encourage patients to commit to using telemedicine visits?”* (Process) (full survey available on request).

Following the completion of surveys, we invited providers meeting the inclusion criteria to participate in semi-structured interviews to ascertain more in-depth feedback regarding their experience in implementation of obstetric telemedicine. A total of 13 providers, some of whom previously completed the anonymous online survey, participated in the in-depth interviews. These interviews consisted of questions identical to those in the online survey, with opportunity to expand upon answers. Interviews lasted approximately 20 to 30 minutes and were audio-recorded and transcribed for data analysis. To preserve confidentiality, each transcript was assigned a pseudonym. This study was reviewed and determined to be exempt as program evaluation by the University of South Florida Institutional Review Board.

### Data Analysis

We developed an initial codebook based on *a priori* structural codes guided by CFIR. This codebook was revised for clarity and the relevance of codes and definitions. Two evaluation team members independently coded four transcripts to consensus (C.R. and K.F.), and a kappa statistic of 0.8 was calculated, indicating good agreement between coders.^18^ Any coding discrepancies were identified and discussed to achieve agreement on the final codebook. One coder (C.R.) coded the remaining surveys and transcripts. A thematic analysis approach was used to analyze the data^19^ and all coding and data analysis was conducted using MAXQDA 2020 software.^20^ Recruitment of providers was ended once it was determined that data saturation was achieved; participant comments were repeated from previous surveys/interviews and no new information was being added. Additionally, trustworthiness was enhanced by conducting peer debriefing (credibility), having two evaluation team members independently code at least ten percent of the interviews with a good kappa (reliability), using provider quotes to represent themes (confirmability), and by having audit trails throughout the process.^21,22^

## Results

The evaluation results from survey and interview participants (Table 1) are summarized below along with representative quotes found in Table 2 (quote ID noted in parentheses), These findings are aligned with CFIR constructs that help to inform the pros and cons of obstetric telemedicine implementation from the viewpoints of a diverse range of providers who are newly conducting this practice.

**Table 2.**
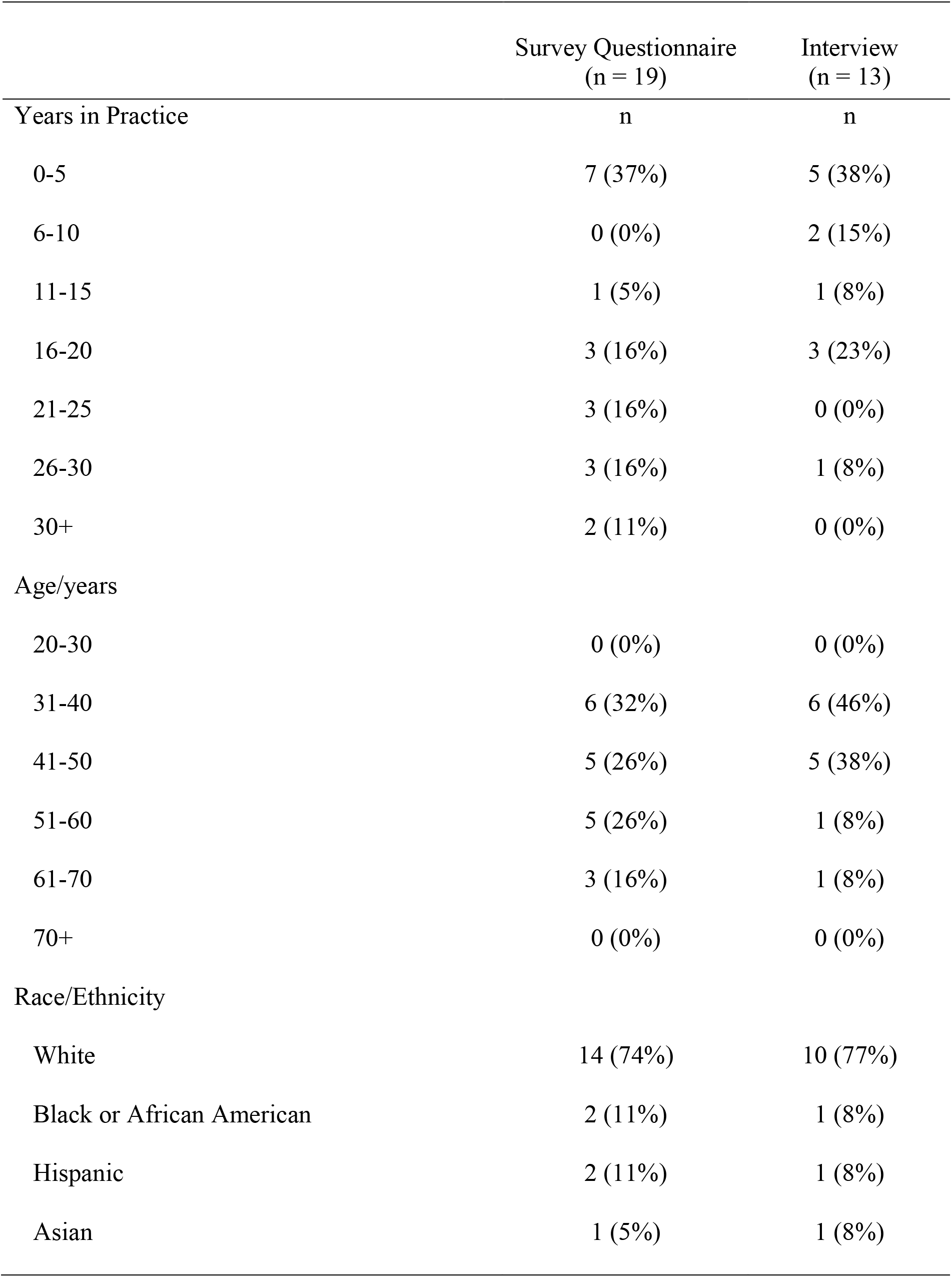
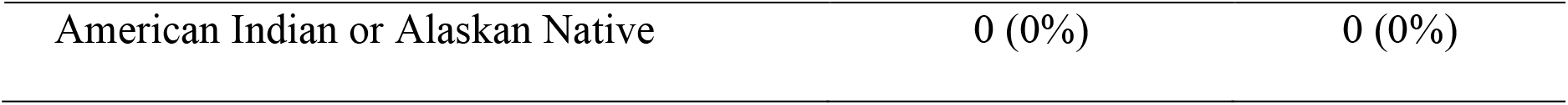
Demographic characteristics of obstetric providers implementing telemedicine

**Table 3.**
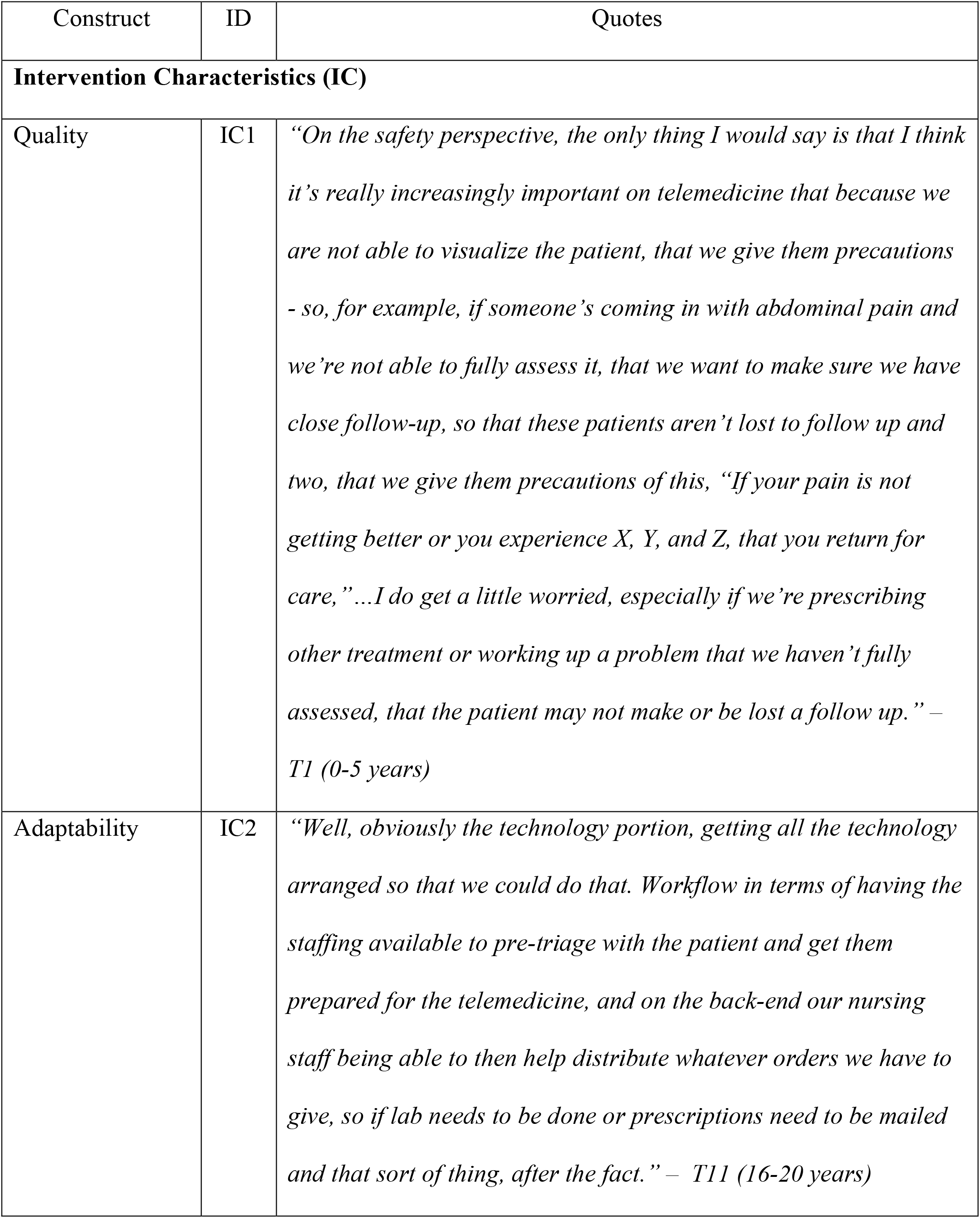
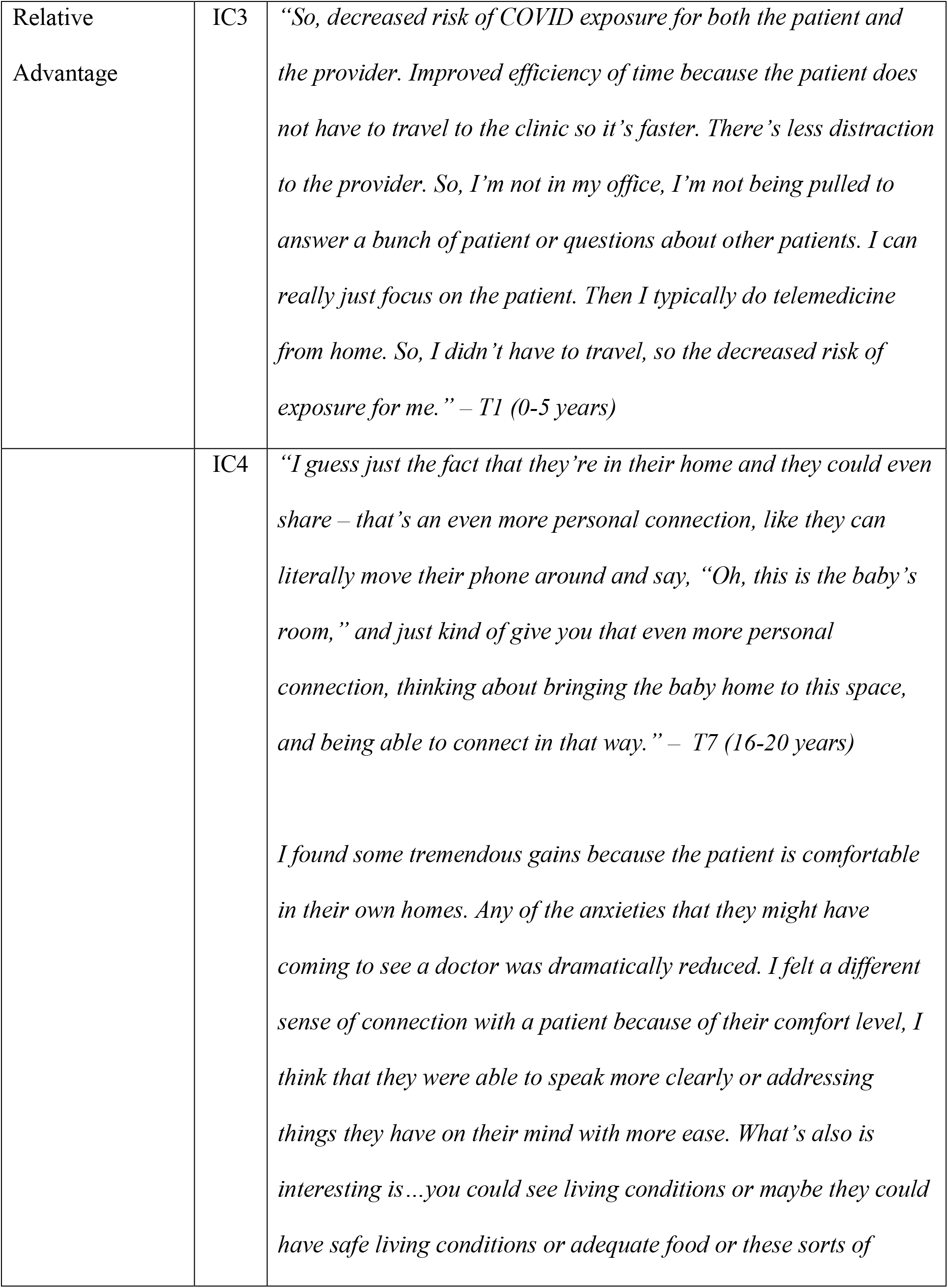
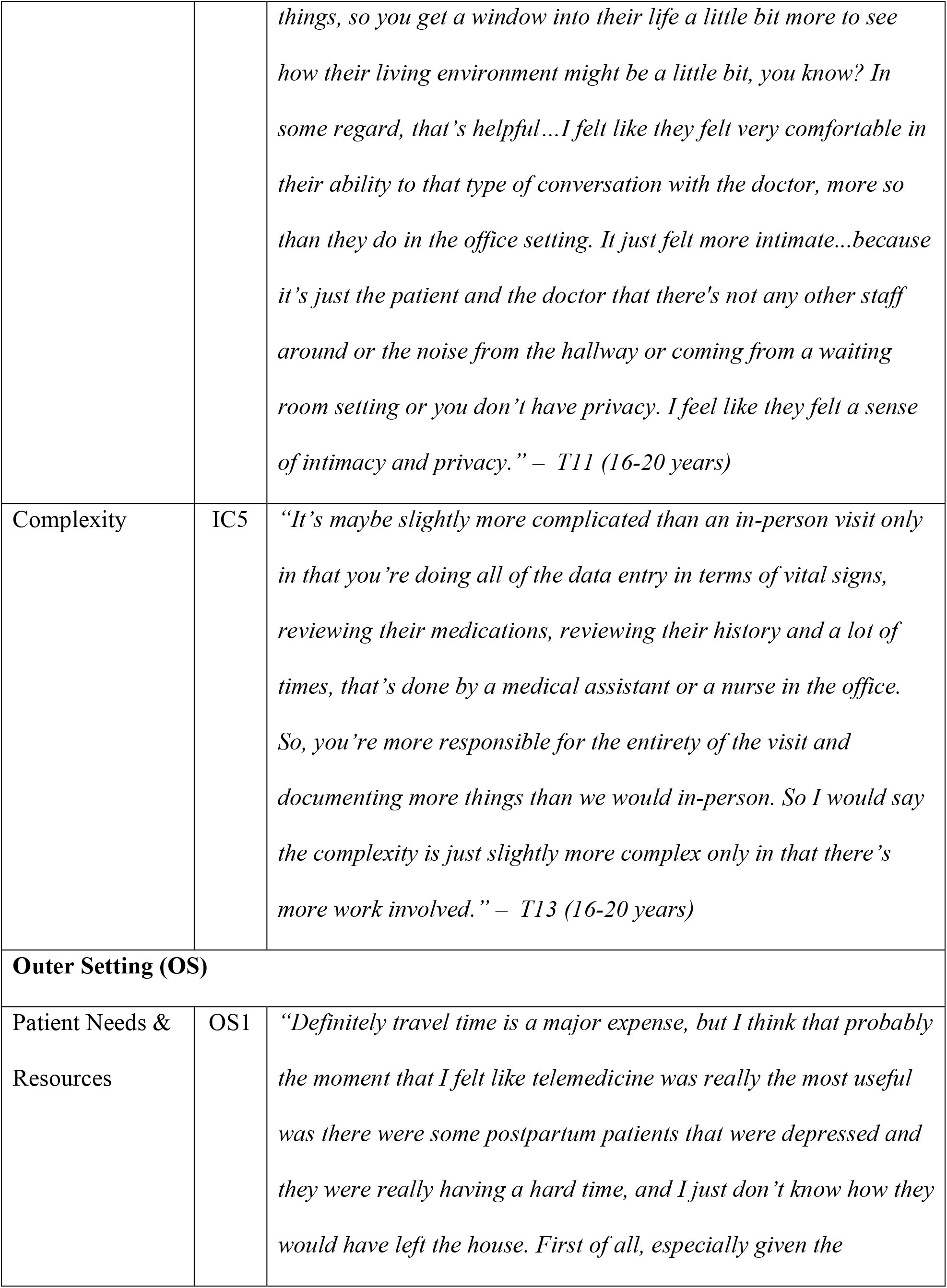
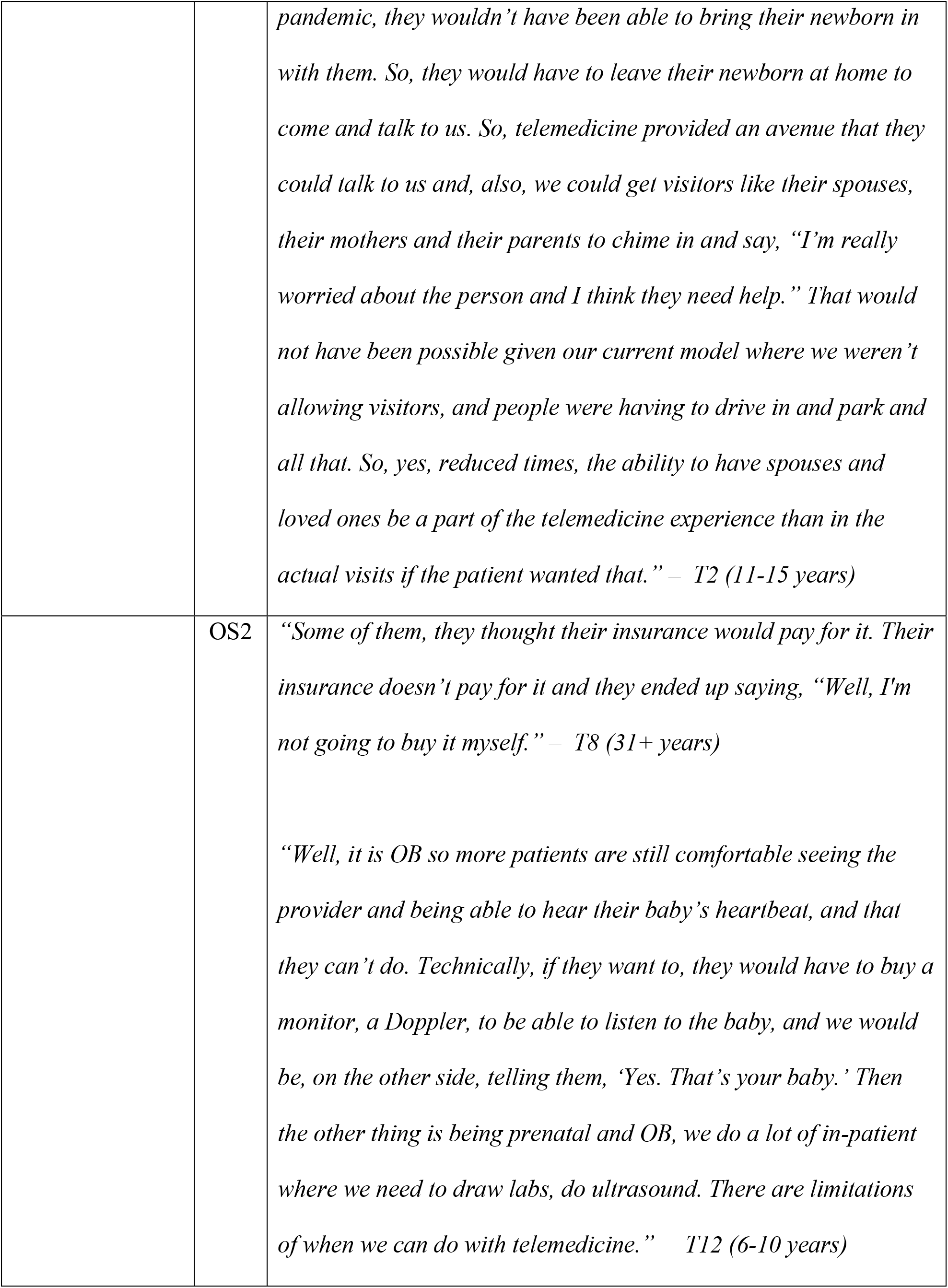
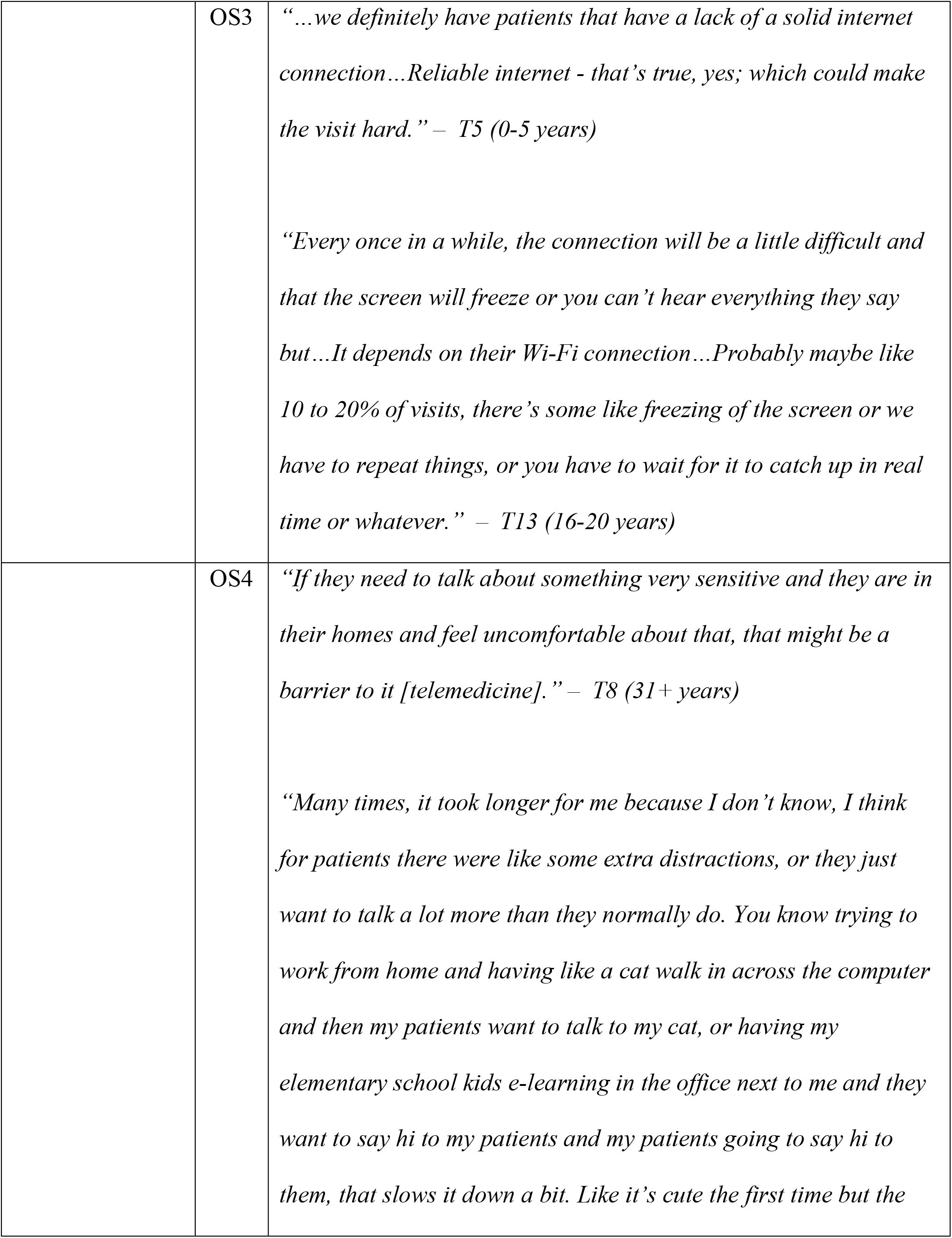
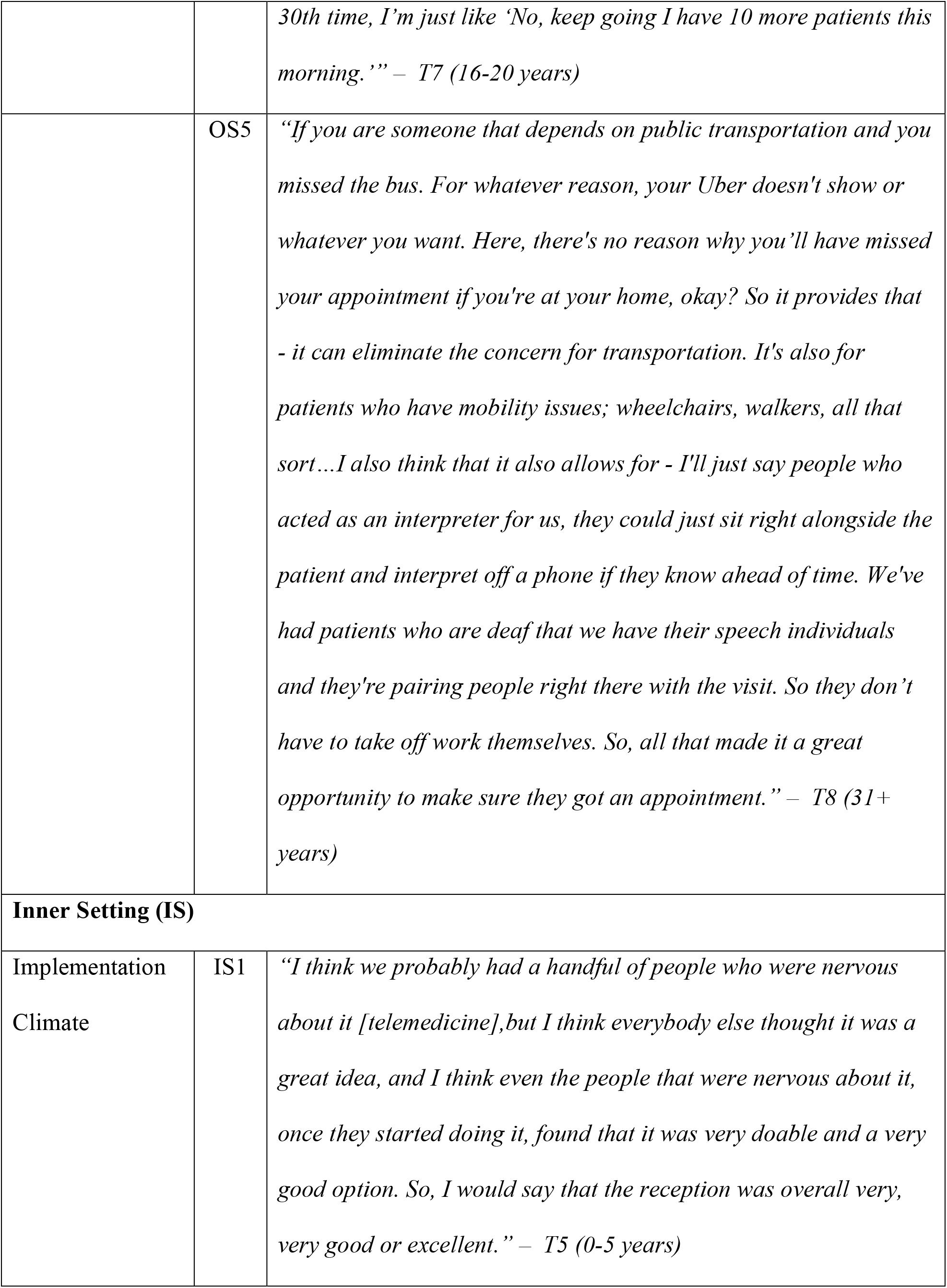
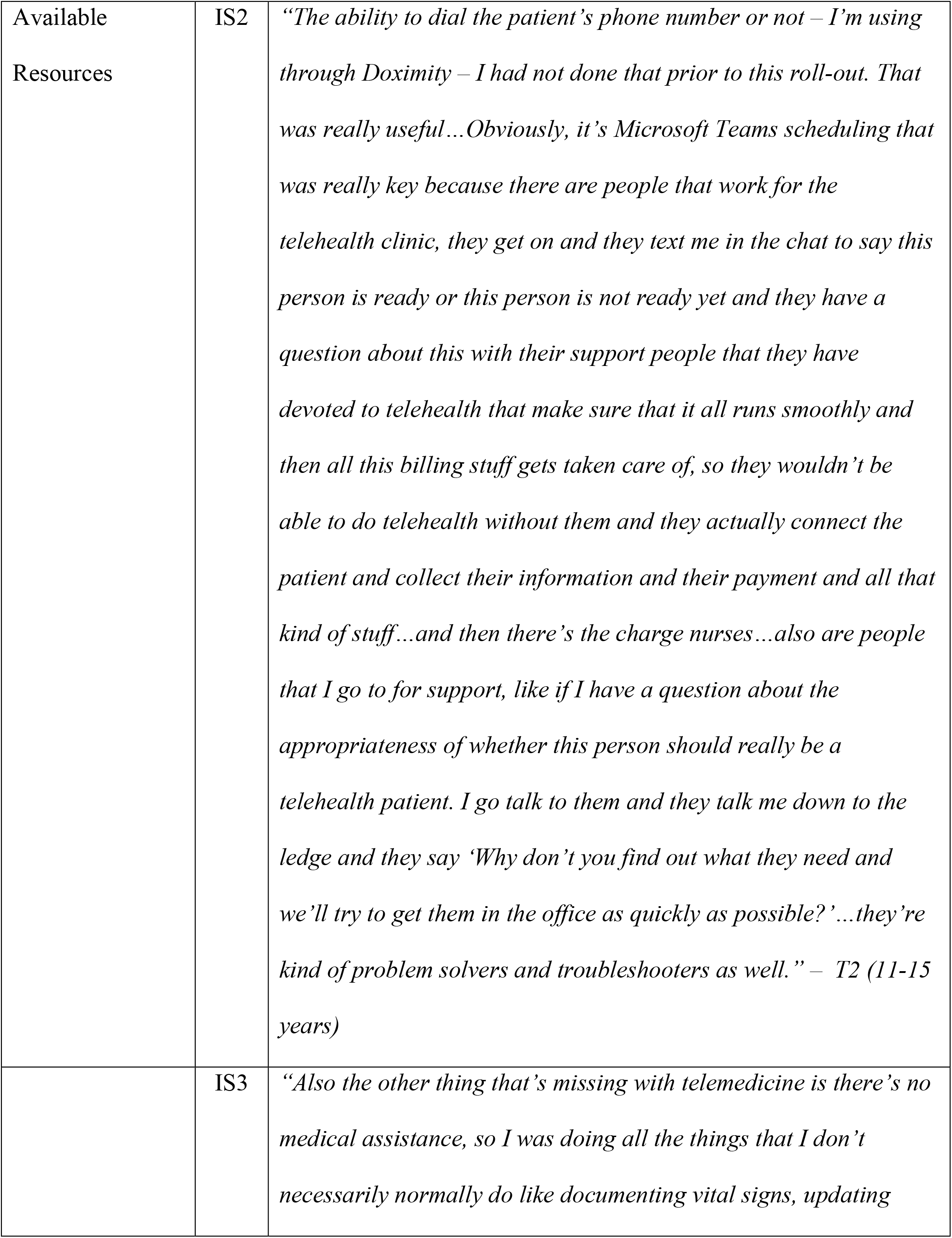
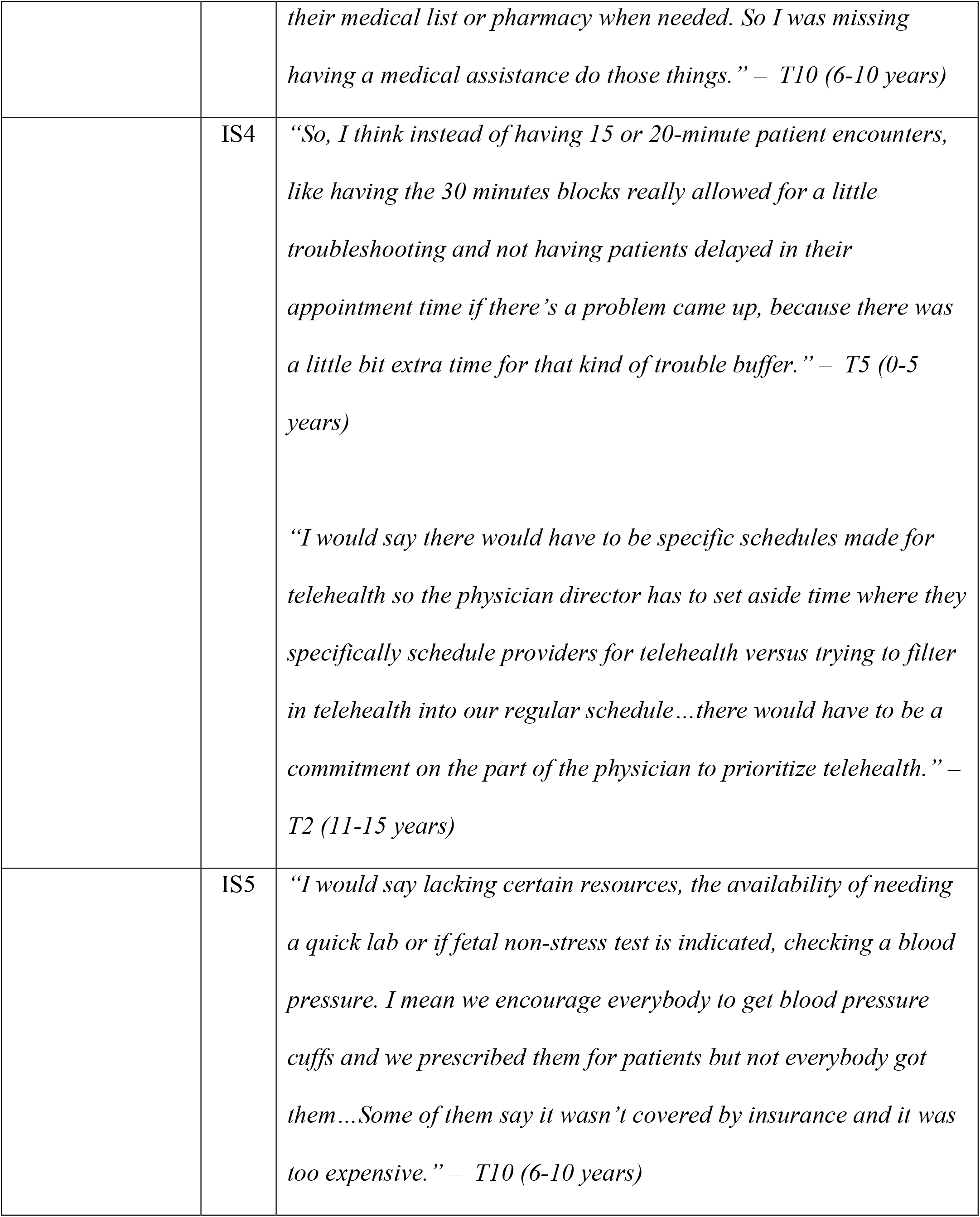
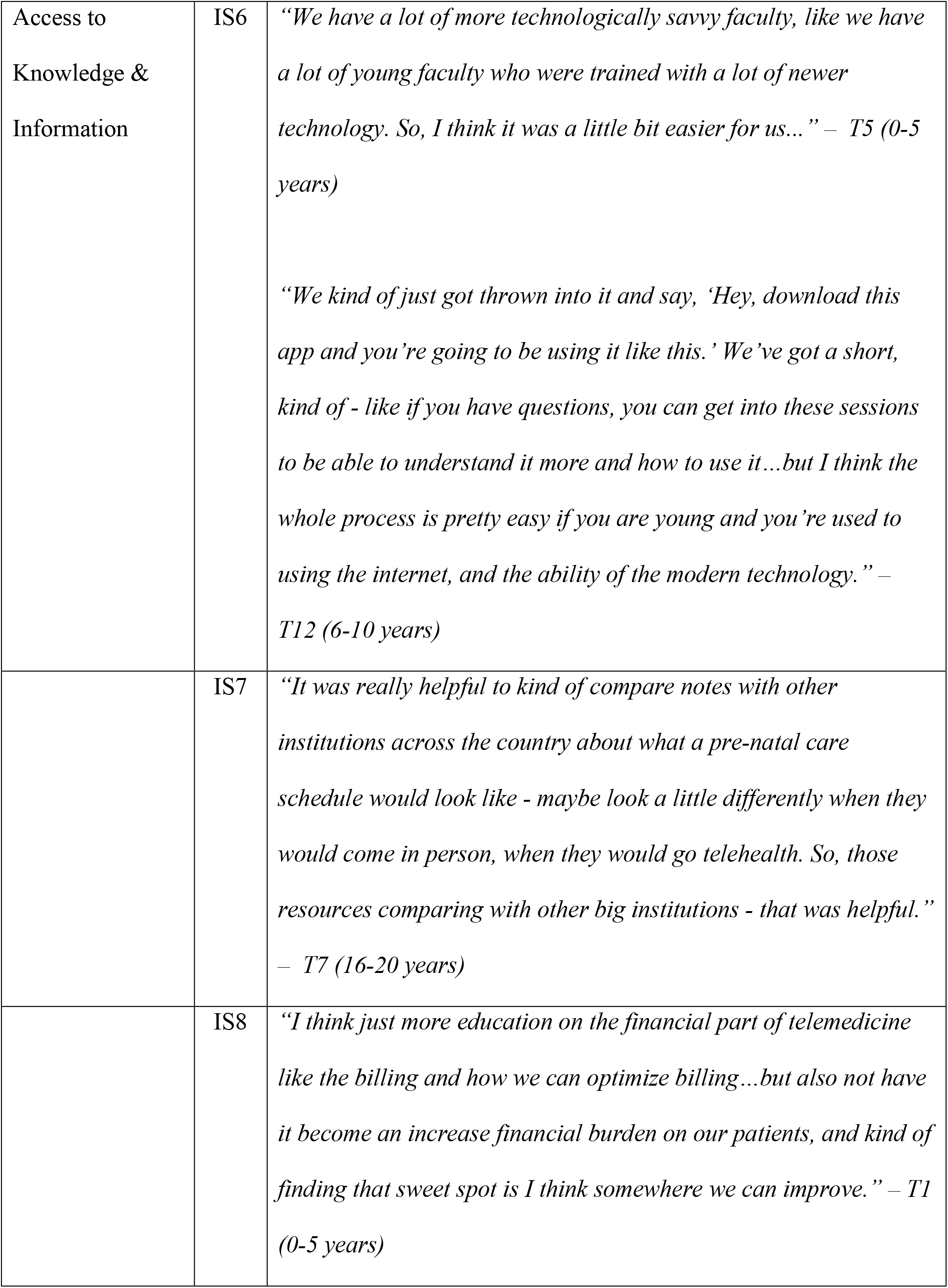
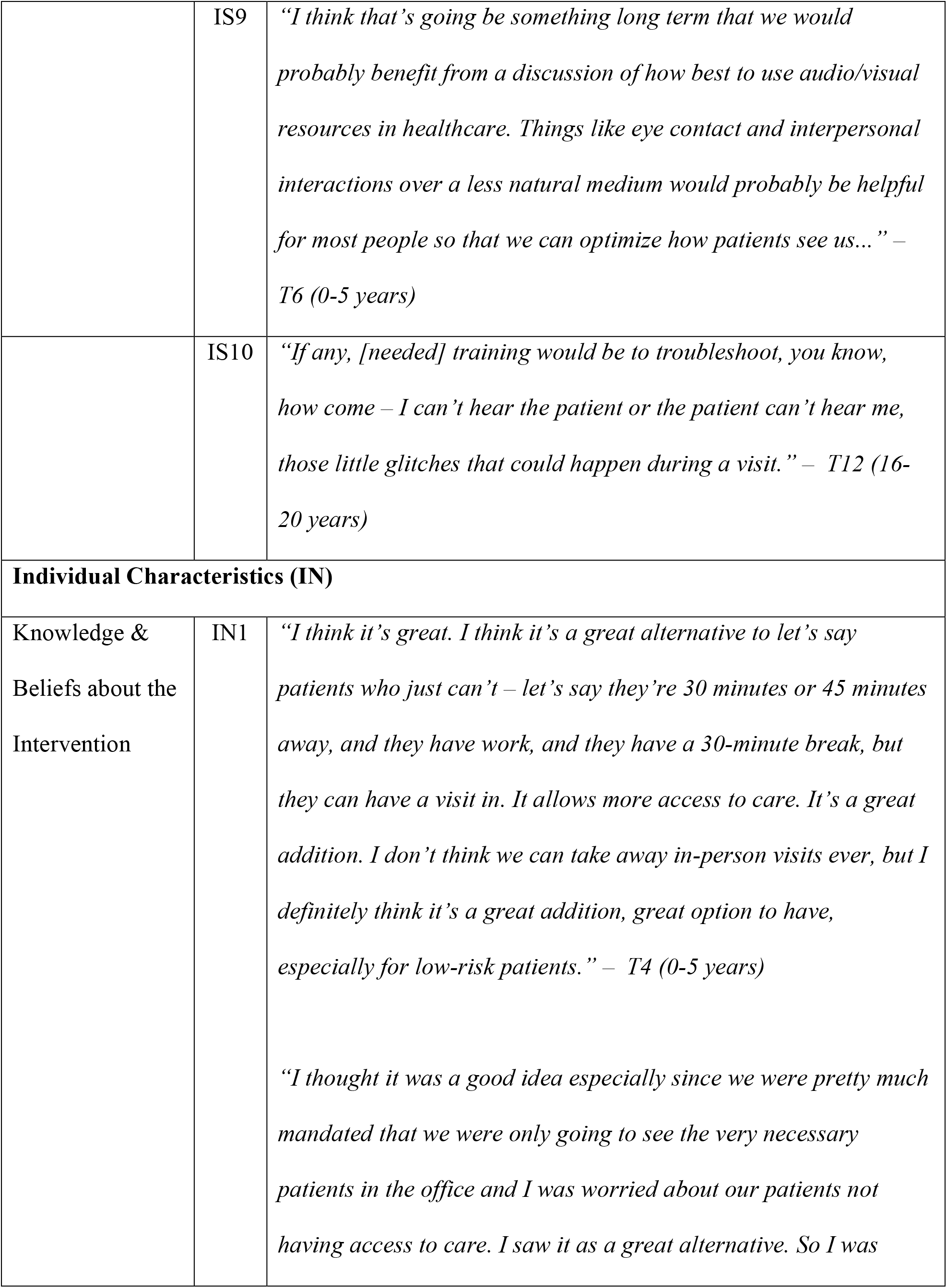
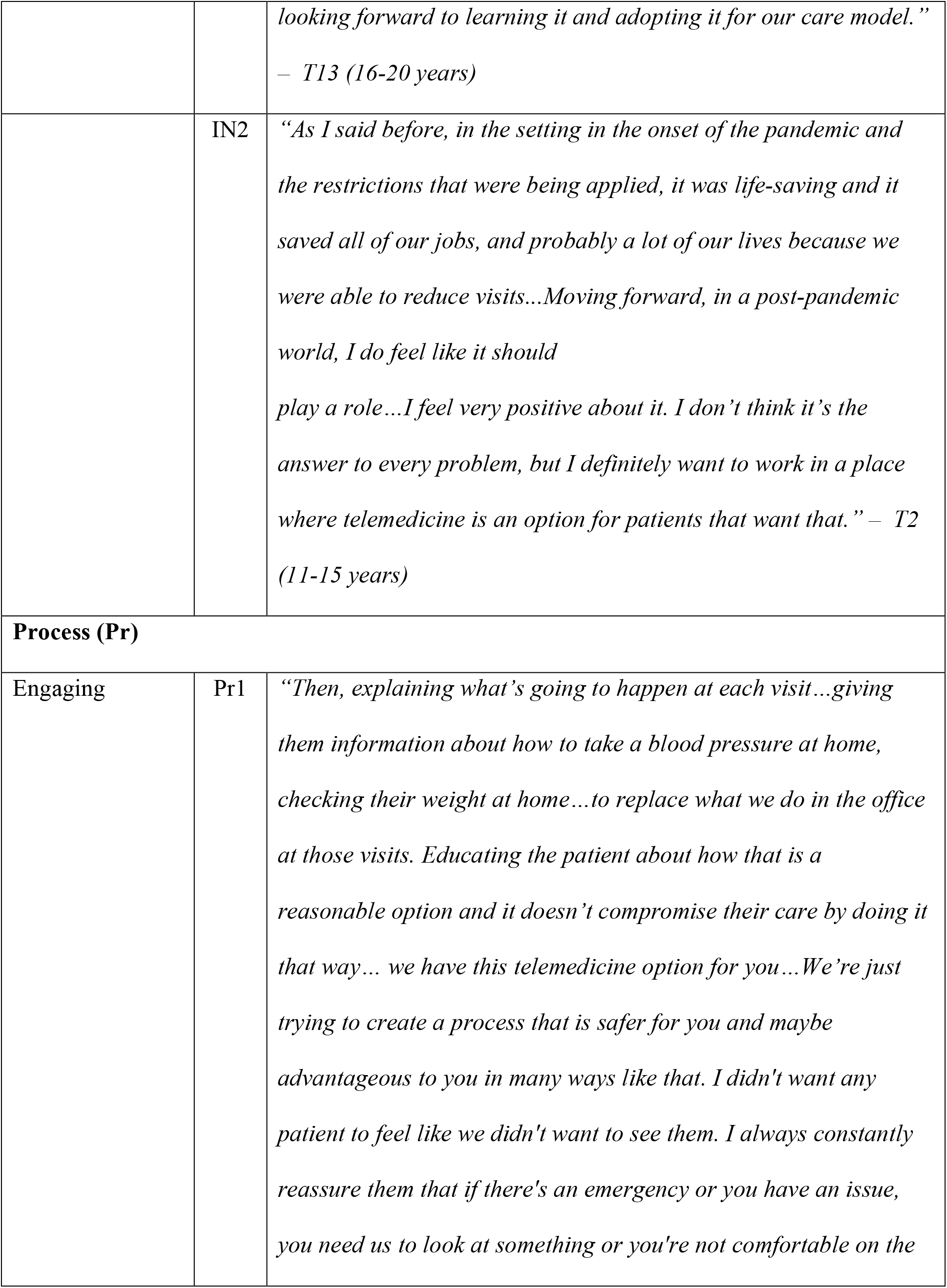
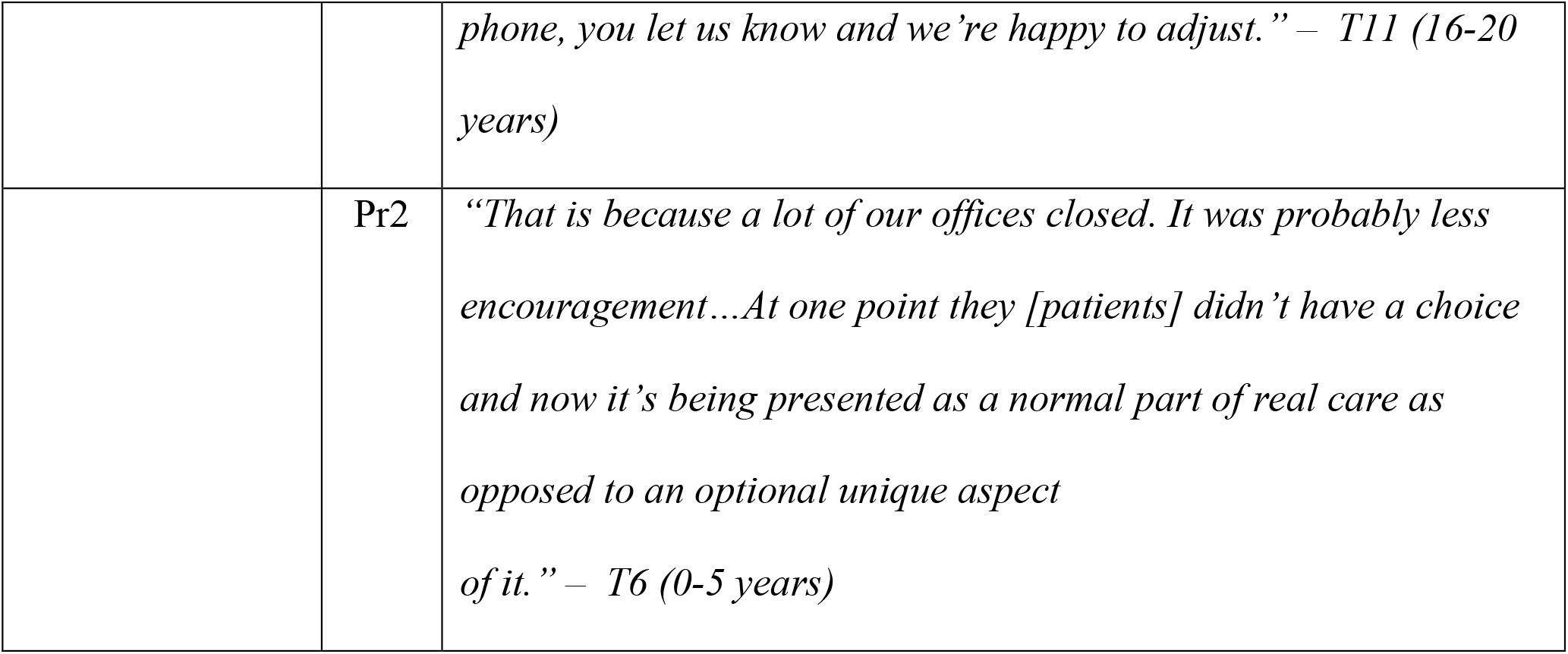
Participant quotes

### Provider description

The participants in this study were predominantly White (74%), with Black (11%), Hispanic (11%) and Asian (5%) providers represented as well (Table 1). Over a third (37%) had been in practice for up to 5 years, while over 58% had been in practice for more than 15 years.

### Provider perceptions of implementation

Prior to this implementation of obstetric telemedicine, almost all providers had never used telemedicine in practice before. With the rapid implementation of obstetric telemedicine early in the COVID-19 pandemic, obstetric providers shared their perceptions.

#### Quality

The quality of telemedicine visits referred to providers’ perception of the safety (avoiding risk or harm) and efficacy of telemedicine visits to have desired outcomes. Most providers did not have any concerns about the safety and efficacy of telemedicine visits. However, a few providers (11 years or more in practice) had concerns about solely using telemedicine for new patients with unrecognized urgent conditions or for patients who required a physical examination or ultrasound (IC1).

#### Adaptability

The adaptability of telemedicine visits entailed the degree to which changes could be made to telemedicine visits for it to work effectively in a clinic. Such changes included the use of multiple platforms (e.g. Microsoft Teams, Doximity dialer, etc.) to conduct telemedicine visits efficiently, the flexibility of both scheduling patient appointments and allotting extra time to transition between scheduled in-person and telemedicine appointments, or simply for troubleshooting technical issues if necessary. Likewise, the ability to incorporate support staff during telemedicine visits to improve workflow made telemedicine work effectively (IC2).

#### Relative Advantage

The relative advantage of telemedicine visits referred to providers’ perception of the advantages of implementing obstetric telemedicine compared to in-person obstetric visits. Generally, providers regarded telemedicine as comparable to in-person obstetric visits. Convenience, comfort, and decreased exposure to COVID-19 were the most common themes associated with the relative advantage of obstetric telemedicine. Convenience was described by providers as less travel commute and parking issues, less visit wait times, reduced need for childcare, fewer missed or late appointments, more flexibility in scheduling appointments, and longer visits for counseling. Additionally, providers expressed added benefits of telemedicine for both patients and providers to include the convenience of less distractions and a decreased risk of exposure to COVID-19 infection (IC3). Providers also discussed the personal connections they were able to have with patients because of how comfortable patients were in their home environment during telemedicine visits (IC4). Although providers described telemedicine as having several advantages compared to traditional in-person visits, most providers agreed that the limitations of not being able to perform physical exams, including assessing vital signs (e.g. blood pressure (BP), fetal heart sounds, weight), patients’ lack of access to labs and ultrasounds, and technology-related issues, made telemedicine less advantageous to in-person visits.

#### Complexity

The complexity of telemedicine visits was defined as the perceived difficulty of its implementation. Most providers described telemedicine visits as simple or not complicated to implement. However, a few providers in practice for 16 years or longer, found telemedicine to be slightly more complex in terms of the multiple steps involved in the process of conducting a telemedicine visit (IC5).

#### Patient Needs & Resources

This construct encompassed the extent to which patient needs, and barriers and facilitators to meet those needs were known and prioritized by the clinic. In the context of the COVID-19 pandemic which restricted access to healthcare services, telemedicine offered an alternative option to access obstetric care. Themes encompassing this construct – patients’ need for convenience, comfort, decreased exposure to COVID-19 infection, and physical examination, were similar to those under the relative advantage construct. For example, one provider discussed how the convenience of obstetric telemedicine visits and the flexibility to include family members for support at these visits was helpful to patients (OS1). Many providers mentioned the need expressed by patients to hear the fetus’ heartbeat during visits. However, this was only possible if the patient had an at-home fetal heart doppler machine.

Barriers to meeting these needs included a lack of patient access to at-home monitoring devices such as BP machines, weight scales, and fetal heart doppler machines to measure vital signs; health insurance policies not covering the purchase of these at-home monitoring devices; the impracticality of having a physical examination, lab work or ultrasound done if required (OS2); and access to technology such as reliable internet service which was required to participate in telemedicine visits (OS3). Some providers mentioned patient privacy and distractions as barriers to patient comfort and a more efficient telemedicine visit (OS4). Overall, providers felt that telemedicine as an alternative option to in-person obstetric visits met most of these patient needs. This was particularly for low-risk obstetric patients not requiring a physical examination and for patients with certain hearing or mobility limitations (OS5).

#### Implementation Climate

Implementation climate referred to the shared receptivity of providers to telemedicine visits within their clinics. Overall, providers were receptive to the implementation of telemedicine visits. One provider reported that although some providers were initially nervous about the implementation of telemedicine, it was however well received (IS1).

#### Readiness for Implementation

The tangible and immediate indicators of overall commitment to the decision to implement telemedicine visits is described by the readiness for implementation construct. Regarding the question *“Did you feel the training prepared you to carry out telemedicine visits?”*, all providers acknowledged feeling prepared to implement telemedicine visits. We further investigated two of the three sub-constructs under the main construct readiness for implementation, that is, 1) available resources and 2) access to knowledge and information.

#### Available Resources

This sub-construct included resources devoted for the implementation and on-going operation of telemedicine visits. Generally, themes associated with resources available for implementation as reported by providers included technology access and support, staff support, time availability, and access to at-home monitoring devices (e.g. BP machine, etc.). For some providers, resources for telemedicine implementation was considered adequate, while for others the opposite held true. For example, access to telemedicine platforms such as Microsoft Teams and Doximity, and the availability of staff to troubleshoot both technical issues and assist with the telemedicine visit process was considered ample (IS2). However, some providers did not have adequate staff support (IS3). Likewise, provision of time allotted for telemedicine visits was sufficient and appropriate for some, but not all providers (IS4). Finally, some patients had access to BP machines, weight scales, and fetal heart dopplers provided to them by the clinic, while other patients did not have access because they were not covered by insurance and were too expensive (IS5). This created a barrier for patients’ desire to hear the baby’s heartbeat and for providers to complete comprehensive telemedicine visits.

#### Access to Knowledge and Information

This construct included access to available information and knowledge associated with OB telemedicine and its use. Providers reported generally gaining knowledge and access to information about telemedicine visits through in-person and online trainings. In particular, providers with fewer years in practice revealed that the process of familiarizing oneself with telemedicine was easier for younger or more tech savvy colleagues (IS6). One provider sought obstetric telemedicine information provided by their clinic and compared it to information provided at other larger institutions (IS7). Although most providers gained knowledge and information from trainings offered, some indicated there was no need for continued trainings, while others indicated certain aspects they considered missing or needing additional trainings. These included information and trainings related to telemedicine billing and reimbursements (IS8), improvements in provider-patient interaction (IS9), and troubleshooting technological issues (IS10).

#### Knowledge & Beliefs about the Intervention

The construct of knowledge and beliefs about obstetric telemedicine referred to providers’ attitudes and values placed towards telemedicine visits and its implementation in their clinics. Overall, all providers had positive reactions to implementation as it related to their knowledge and beliefs about telemedicine. Providers mostly considered telemedicine a usefulness alternative to deliver obstetric care especially for low-risk obstetric patients (IN1). Additionally, some providers believed the implementation of telemedicine enabled obstetric providers to continue employment during the COVID-19 pandemic, and despite few providers believing its implementation was rushed, some providers expressed the desire for telemedicine to continue beyond the pandemic (IN2).

#### Engaging

The question *“What steps were taken to encourage patients to commit to using telemedicine visits?”* referred to the construct of engaging. Mostly, providers reported that patients were encouraged to commit to participating in telemedicine visits by educating them about the simplicity and advantages of telemedicine (similar to those mentioned under the relative advantage construct), explaining to them what the process of a typical telemedicine visit entailed, as well as reassuring them of adequate patient care (Pr1). One provider reported that because of the COVID-19 pandemic, patients were not given an option to opt-out of telemedicine visits. Nevertheless, it has now become part of routine care in the clinic (Pr2).

## Discussion

The implementation of the obstetric telemedicine care model was deemed favorable, safe and an alternative option for patients during the COVID-19 pandemic. The majority of providers found this model of care to be easy to use and implement even though they did not have past exposure to telemedicine. Barriers to telemedicine noted included lack of home monitoring devices for blood pressure and fetal heart tones, knowledge of billing, and concerns regarding patient privacy. These advantages and barriers are similar to those reported in other studies analyzing rapid telemedicine implementation in the post-COVID era.^8–14^

It is encouraging that most of the barriers identified by providers can be addressed. Furthermore, the pros noted in this evaluation suggest that it may be advantageous to extend telemedicine beyond the pandemic in order to improve the efficiency and access of healthcare services provision in the future. The American College of Obstetricians and Gynecologists has provided guidance to providers in the form of a telemedicine implementation guide^23^ and guide for telemedicine implementation during the pandemic.^24^

### Strengths and Limitations

One of the major strengths of this study was the ability to quickly evaluate a rapid implementation of telemedicine during a pandemic. We were able to capture this feedback during the rollout of a new model of care and in a prospective fashion. Our results compare favorably with the current research which shows that telemedicine was well received during the COVID pandemic.^8–14^ Additionally, we were able to perform this evaluation with a robust theoretical framework. Weaknesses include that this study is single site within the context of a unique situation (rapid implementation due to the COVID-19 pandemic); therefore, the results may differ from future implementation of telemedicine where more advanced planning and training would be possible prior to implementation.

### Future research

Future research is needed on how to integrate telemedicine into care for high-risk pregnancies, patient privacy concerns and access to internet and at-home monitoring devices. Additional funding is needed to provide patients with at home monitoring devices through insurance reimbursement or other funding mechanisms, especially for low-income women who cannot afford these devices due to high out-of-pocket costs. Additional guidance and standardization of a telemedicine prenatal care model for low-risk women from national professional organizations such as ACOG and the American College of Nurse-Midwives would be advantageous for both providers and patients in implementation of this model across the country. For high-risk women, more research is needed to determine the best protocols for surveillance and number of visits needed during pregnancy. Further evaluation is also needed to explore this care model from patient perspectives. It would be useful to learn of the strategies used and challenges encountered from the perspectives of leadership/administration, and to further explore the outer setting of the community context related to obstetric care access.

## Conclusion

The implementation of the obstetric telemedicine care model was deemed a favorable, safe alternative option for patients during the COVID-19 pandemic in our program evaluation and shows promise for continuation. Future research and evaluation should assess providers’ and patients’ perspectives about a hybrid obstetric care model, patient privacy concerns and access to internet and at-home monitoring devices.

## Data Availability

De-identified data available only with Data Use Agreement with University of South Florida

